## Supplementary material for "Comparison of pretraining models and strategies for health-related social media text classification"

### Appendix I

Each of RoBERTa, BERTweet, and BioBERT has multiple implementations which differ in the model size, the pre-training data size, and the training time. The specific models for RoBERTa, BERTweet and BioBERT we used in our experiments are RoBERTa_Base, BERTweet_Base and BioBERT v1.1.

### Appendix II

| **Hyper-parameter** | |
|---|---|
| Number of Layers | 12 |
| Hidden size | 768 |
| FFN inner hidden size | 3072 |
| Attention heads | 12 |
| Attention head size | 64 |
| Dropout | 0.1 |
| Attention Dropout | 0.1 |
| Weight Decay | 0.01 |
| Learning Rate Decay | Linear |
| Adam $\beta_1$ | 0.9 |
| Adam $\beta_2$ | 0.98 |
| Gradient Clipping | 0 |
| Batch Size | 32 |
| Warmup Ratio | 0 |
| Adam Epsilon | $1 \times 10^{-8}$ |

**Table 1**. The full hyper-parameter configurations of the classification model. The maximum sequence size of each model is set as the same as the off-the-shelf model, which is 128 for BERTweet and 512 for the rest models.

### Appendix III

| ID | Task | Link |
|---|---|---|
| 1 | ADR Detection | http://diego.asu.edu/Publications/ADRClassify.html |
| 2 | Breast Cancer | https://healthlanguageprocessing.org/smm4h-2021/task-8/<br>Data also available from: https://sarkerlab.org/blogs/smm-bc.html |
| 3 | NPMU characterization | Dataset is currently protected due to the sensitive nature of the posts (based on suggestions by the funding body—National Institute on Drug Abuse). Data will be made available upon request |

| | | |
|---|---|---|
| | | following a data use agreement (DUA). Please email the PI Abeed Sarker at for data access. |
| 4 | WNUT-20-task2 (informative COVID-19 tweet detection) | https://github.com/VinAIResearch/COVID19Tweet |
| 5 | SMM4H-17-task1 (ADR detection) | https://healthlanguageprocessing.org/sharedtask2/ |
| 6 | SMM4H-17-task2 (medication consumption) | |
| 7 | SMM4H-21-task1 (ADR detection) | https://healthlanguageprocessing.org/smm4h-2021/task-1/ |
| 8 | SMM4H-21-task3a (regimen change on Twitter) | https://healthlanguageprocessing.org/smm4h-2021/task-3/ |
| 9 | SMM4H-21-task3b (regimen change on WebMD) | |
| 10 | SMM4H-21-task4 (adverse pregnancy outcomes) | https://healthlanguageprocessing.org/smm4h-2021/task-4/ |
| 11 | SMM4H-21-task5 (COVID-19 potential case) | https://healthlanguageprocessing.org/smm4h-2021/task-5/ |
| 12 | SMM4H-21-task6 (COVID-19 symptom) | https://healthlanguageprocessing.org/smm4h-2021/task-6/ |
| 13 | Suicidal Ideation Detection | https://zenodo.org/record/4278895#.YT_KL1POkWo |
| 14 | Drug Addiction and Recovery Intervention | https://zenodo.org/record/4543776#.YUJW-FPOkWo |
| 15 | eRisk-21-task1 (Signs of Pathological Gambling) | https://early.irlab.org/ |
| 16 | eRisk-21-task2 (Signs of Self-Harm) | |
| 17 | Sentiment Analysis in e-Health Forums (Food Allergy Related) | https://zenodo.org/record/1479354#.YVHTeFPOkWo |
| 18 | Sentiment Analysis in e-Health Forums (Crohn'S Disease Related) | |

| | |
|---|---|
| 19 | Sentiment Analysis in e-Health Forums (Breast Cancer Related) |
| 20 | Factuality Classification in e-Health Forums (Food Allergy Related) |
| 21 | Factuality Classification in e-Health Forums (Crohn'S Disease Related) |
| 22 | Factuality Classification in e-Health Forums (Breast Cancer Related) |

**Table 2**. The links and mechanisms to access the datasets included in this study. For Task 7-12 and Task 15-16, the datasets are not publicly available but can potentially be accessed by sending requests to the shared task organizers.
